# Data-centric Artificial Intelligence and Cancer Research: Construction of a Real-World Head and Neck Treatment Data Repository

**DOI:** 10.1101/2025.02.12.25322092

**Authors:** Victoria Butterworth, Tom Young, Haleema Drake, Isabel Palmer, Tania Avgoulea, Eleanor Ivy, Joshua Andriolo, Carole Creppy, Corla Routledge, Delali Adjogatse, Anthony Kong, Imran Petkar, Miguel Reis Ferreira, David Eaton, Mary Lei, Sarah Misson, Dijana Vilic, Teresa Guerrero Urbano

**Author notes:** Corresponding author: Victoria Butterworth, Radiotherapy Physics, Guy’s Cancer Centre, Great Maze Pond, London, United Kingdom, SE1 9RT.

## Abstract

**Background and Purpose:** The performance and generalisability of machine learning (ML) models relies on high-quality data. Retrospective and prospective collection of high-quality data for research use whilst respecting data protection and patient privacy remains a challenge in the clinical environment. Currently, months of laborious extraction and clinical annotation are often necessary before data analysis can begin. We present a novel institutional federated data lake, utilising open-source software, to facilitate efficient production of ML models from Head and Neck Cancer (HNC) imaging and Radiotherapy (RT) data. This structured pipeline dramatically reduces the time associated with the production of ML models and real-world evidence generation. This paper describes our governance-compliant processes and provides a framework for establishing similar databases.

**Materials and Methods:** XNAT, is a powerful open-source imaging platform. Within our department, it forms a part of the local secure enclave for the purposes of federated learning in artificial intelligence projects and provides import, archiving, processing, search and secure distribution facilities for imaging and RT data.

**Results:** We have created a clinically annotated, carefully curated, data lake of 2,895 consenting HNC patients containing 22,170 relevant diagnostic, staging, treatment and monitoring imaging sets. Key recommendations for replication include infrastructure planning, robust patient and data selection criteria and prioritising patient consent and privacy.

**Conclusions:** This secure and extensible imaging and HNC RT cancer database set-up promises to be an exceedingly useful tool for research, revolutionising the time and cost associated with the production of ML models, making the process safer, faster and more efficient.

**Highlights:**

- Real-world data is critical for building predictive clinical models.
- Curation of clinical data for machine learning can necessitate months of laborious extraction and annotation.
- Patients have the right to have their data handled with the highest standards of respect, security and governance.
- We describe a database infrastructure constructed under a transparent and safe system of control and stewardship.
- This secure and extensible structured database dramatically reduces data extraction time for AI-driven cancer research.

## Background

Cancer remains one of the leading causes of death in the UK and it is estimated that 1 in 2 people will be diagnosed at some point during their lifetime [1]. Head and neck cancers (HNC) are a diverse group of malignancies affecting areas such as the oral cavity, pharynx and larynx [2], and each year over 12,000 new cases are diagnosed in the UK. HNC is more common in men and significant risk factors include tobacco, alcohol and human papillomavirus (HPV) infection [3].The burden of disease is large and, whilst the 10-year survival varies between 19-59% [1], the side effects from treatment can result in significant morbidity, with significant to cost to healthcare systems.

Radiotherapy (RT) is a key treatment of HNC [2], often delivered with curative intent, but many patients experience recurrence, progression or significant long-term toxicities despite advances in imaging, planning and treatment delivery. Predictive models integrating outcome and RT data underpin many of the dose-volume constraints used in clinical practice to limit doses to organs at risk. Models have also been developed to try and predict tumour control outcomes. These models, however, vary in their level of complexity, many lack validation, and most are not used in the clinic. Recently, machine learning (ML) has been proposed to improve on previous models. This is because it provides the potential to model multifactorial RT outcomes, can incorporate RT imaging (i.e. dose maps) in addition to the DVH and fractionation data used by traditional modelling, is able to consider multiple clinical and biological prognostic factors and can use large clinical datasets [4][5][6]. The resulting improved predictive models could enable individualized RT with the aim to increase tumour control whilst reducing radiation toxicity and improve patients’ quality of life.

Randomized Clinical Trials (RCTs) provide the highest level of evidence for treatment efficacy but stringent eligibility criteria can create selection biases resulting in real world population outcomes not matching those of RCTs [7]. Analysis of real-world data (RWE) can also help answer relevant clinical and policy questions unable to be directly answered using RCTs [8] including long-term follow-ups [9]. RWE can describe treatment outcomes from a more heterogeneous population, in particular through the inclusion of under-represented groups, i.e. due to age and/or those excluded i.e. due to certain co-morbidities. Alternative data collection methodologies include longitudinal observation studies, registry based clinical trials and prospective databases [10].

ML-based predictive modelling requires extensive structured clinical and RT data. Currently clinical data is not stored in a research-ready format and therefore ML model development necessitates months of laborious extraction and annotation [11]. We aimed to establish data infrastructure to enable meaningful and rapid scientific advances [12][13] whilst preserving the integrity of patient consent.

A database of clinically annotated diagnostic imaging and RT treatment data that can be readily accessible and usable under a transparent and safe system of governance and stewardship as well as patient consent and ethics approval has been constructed adhering to the FAIR guiding principles regarding the use and reuse of digital data [14]. This paper describes our experience of setting up the retrospective cohort of the database and recommendations for technical and practical considerations to construct a valuable research tool.

## Methods

### Data lake infrastructure

Extensible NeuroImaging Archival Toolkit (XNAT) [15] is a powerful open-source platform for storing and managing medical imaging and associated clinical data, offering import, archiving, processing and secure distribution facilities. Originally designed for neuroimaging DICOM data, it now supports RT data sharing both nationally [16] and internationally [17].

Within our organisation, XNAT forms part of the local secure enclave for bulk data extraction and machine learning [18]. Independent of electronic patient record provider and RT vendor, it enables the incorporation of legacy RT as well as diagnostic imaging data. A specific Head and Neck XNAT (HN-XNAT) project within the Trust’s secure environment holds identifiable patient data enabling datasets to be updated with new events. Access is restricted to administrators.

XNAT is connected to Sectra PACS (Sectra AB, Linköping, Sweden) via a DICOM node, allowing DICOM query/retrieve (Q/R) to retrieve imaging data stored within PACS. A DICOM Q/R service was set up on our current treatment planning system (TPS), Eclipse (Varian Medical Systems, Palo Alto, CA, USA) using the Database Service option. This enabled the retrieval of RT images, structure sets, dose, RT plans, any additional imaging and spatial registrations used during treatment planning. For patients treated through record and verify (R&V) system Aria (Varian Medical Systems, Palo Alto, CA, USA), the on-set imaging, cone beam CTs (CBCTs) and RT Beams Treatment Record information were also available for retrieval.

In addition, a secondary RT-XNAT (HNRT-XNAT) has been established locally within the RT department to host anonymised data for approved research projects. Only users related to each project are given access to their data and only administrators have full view of all data available, thus complying with best IG practice and data minimisation principles. All database users undergo additional training hosted by the Trust’s Clinical Scientific Computing group in data security. A simplified network diagram of this set up is displayed in Figure 1. A link is also established with our partner university to allow cross-institution anonymised data transfer for projects subject to appropriate governance approvals, including data sharing agreements.

**Figure 1:**
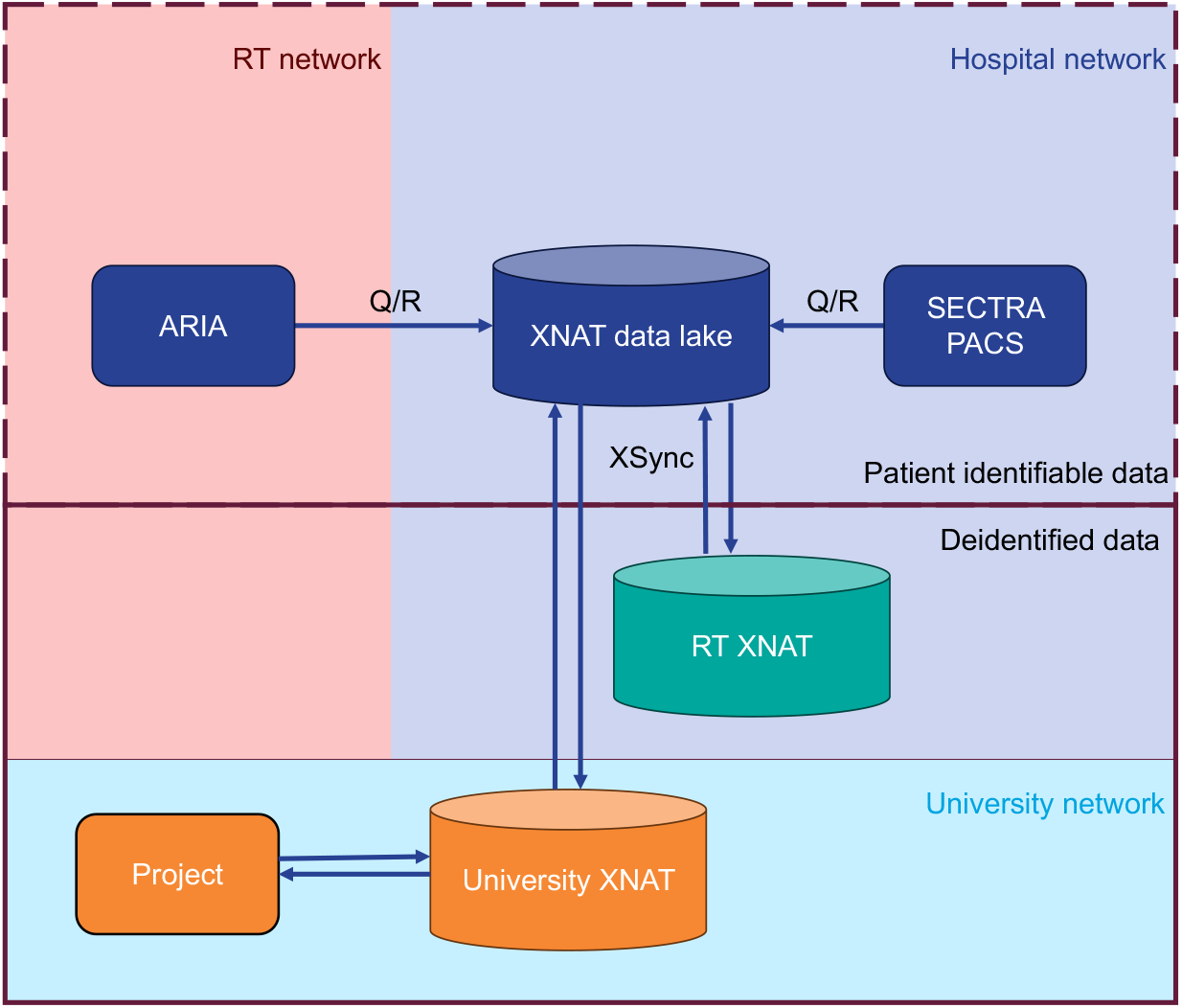
Simplified network diagram of XNAT data lake infrastructure and RT XNAT for research data processing.

### Governance

The project was discussed extensively with the local Information Governance team, which approved storing unanonymised data in the secure on-site data lake with controlled access. All necessary governance approvals including system security policies were obtained.

Clinical audit, quality improvement and service evaluation projects follow local trust policy on data access. Patients are informed their personal health information will be used for these purposes via patient information materials which describe the processes and contribution to the quality and safety of patient care. Audits and service evaluations are registered locally by the Trust.

Any research use of the data requires approval by the Research Ethics Committee (REC) as per national NHS policy [19]. The Guy’s Cancer Cohort (GCC) is a valuable pre-existing REC approved research framework (Reference: 18/NW/0297, IRAS Project ID: 231443) [20] for the use of clinical data. All adult patients are eligible following their first visit for a diagnosis of cancer and they can opt-out of inclusion at any point in their treatment pathway. Opt-out rates are very low, which has been supported by qualitative research on patient preferences regarding opt out consent in our patient population [21]. Patients are provided with an Information Governance approved NHS document that explains how their routinely collected clinical data may be used for research in an anonymised fashion. The document explains the information held, how it is kept safe and accurate, how it supports direct care and medical research and how they can opt out of inclusion within the cohort, ensuring informed consent is gained from each individual. A scientific access committee reviews applications to access the data based on scientific merit, study design and the applicant’s resources. Retrospective clinical data dating back to 2005 is permitted to be utilised as per REC approval [20].

No data leaves the data lake without explicit approval via an audit project ID or GCC approval. The Radiotherapy Development Group assesses project feasibility, while the HN-XNAT Research Access Panel provides investigators with anonymised HNRT-XNAT data as well as access to model production platforms via the Clinical Scientific Computing group if required [22].

### Patient Selection

HNC patients seen by the clinical oncology H&N team between the introduction of inversely planned intensity modulated radiotherapy (IMRT) for radical RT patients (March 2011) and the introduction of a new hospital-wide electronic health record (EHR) system, Epic (Epic Systems Corporation, Verona, WI, USA), in October 2023, were included within the initial cohort.

Patients with skin primaries, paraganglioma and thyroid were excluded, in addition to patients who went on to have their treatment elsewhere and patients treated under different teams. Patients were assigned a single globally unique and persistent identifier [14], their NHS number, to align data sources and prevent record duplication. Patients without NHS numbers (n=8) received a separate unique identifier [14].

### Data Selection

#### Imaging data

PACS was queried for study descriptions and accession numbers of all diagnostic imaging sessions per patient. Relevant imaging studies were selected using study descriptions to ensure only relevant data was ingested via the REST-API, which automated data retrieval, into the data lake. Relevant studies included all orders pertinent to the diagnosis, staging, progression or ongoing monitoring of treatment side effects of HNC. This included all staging ^18^F-FDG PET-CTs, staging and anatomical (head, neck, chest, abdomen and pelvis) CTs/MRIs and all dental x-rays (XR) and video fluoroscopy (VF) studies. Unspecified CTs were inspected and included if relevant. The data flow diagram can be viewed in supplementary material Figure S.1.

#### Radiotherapy data

RT treatment records for patients treated at our institution were retrieved and stored in a .csv file attached to the XNAT data lake. Over the time period of data collection there were 3 TPS, 2 R&V systems, 3 locations of on-set imaging storage and numerous incremental improvements to treatment planning techniques. In 2011, inversely planned IMRT was produced using Monaco TPS (Elekta AB, Stockholm, Sweden), delivered with Mosaiq (Elekta AB, Stockholm, Sweden) as the R&V system and imaged with CBCTs using XVI (Elekta AB, Stockholm, Sweden). 3D-conformal radiotherapy (3D-CRT) plans for palliative patients or simpler patient plans were constructed in XiO (Elekta AB, Stockholm, Sweden) and delivered through Mosaiq. In May 2017, the HNC RT service moved to a new centre and Eclipse (Varian Medical Systems, Palo Alto, CA, USA) was used to create all patient treatment plans with Mosaiq as the R&V system and Mosaiq Data Director (Elekta AB, Stockholm, Sweden) was used to store patient imaging. In October 2021, the R&V system was moved to Aria (Varian Medical Systems, Palo Alto, CA, USA) with imaging stored here also.

Due to the time, costs and difficulties associated with retrieving legacy RT data stored within proprietary file formats, decisions were made as to the inclusion of radiotherapy data based on the relevance of the treatment to modern and current treatment techniques, the ease of data retrieval and the paucity of the data. For this reason, only patients treated using IMRT and VMAT were retrieved from the legacy TPSs. This led to the exclusion of all palliative RT datasets before 2017 as well as cases treated with 3D-CRT. Similarly, CBCTs from before October 2021 were not retrieved. The data retrieved for each patient cohort is summarised in supplementary material Figure S.2. The relevant legacy treatment plans were imported into the current TPS to facilitate ease of treatment planning should the patients return for additional radiotherapy, and to enable a one-step transfer process of all RT data into XNAT.

Available radiotherapy planning CTs, additional imaging, structure sets, plans and dose cubes and on-set imaging were retrieved and ingested into XNAT. The data flow diagram can be viewed in supplementary material Figure S.3. All original and replanned RT data were included, with fractions annotated for accurate future dose data mining.

#### Clinical data

Clinical annotation was built using multiple sources and methods to ensure completeness and accuracy. Structured data extraction for the identified 2985 patients used the Health Catalyst platform (Health Catalyst Inc., Salt Lake City, UT, USA), to consolidate clinical data from legacy electronic health record systems into an Enterprise Data Warehouse (EDW). Structured Query Language queries enabled extraction of data tables containing demographic, disease, treatment, and outcome data. Unstructured data was mined using CogStack [23], a natural language processing tool, which identified previously validated SNOMED CT concepts within the clinical documents. Previously existing research datasets and histopathology records were integrated into the EDW with limited targeted manual curation supplementing missing information. Rigorous validation processes ensured the dataset met established quality standards [24].

## Results

### Data Lake

We have created a clinically annotated, carefully curated, data lake of medical imaging data of 2895 consenting H&N patients for the purposes of federated machine learning containing over 22170 relevant clinical diagnostic, staging, treatment and monitoring imaging sets.

### Data Access Pipeline

A research access pipeline was created adhering to the detailed governance procedures previously described. Figure 2 shows the diagram detailing data controllers at each stage of the research access pipeline.

**Figure 2:**
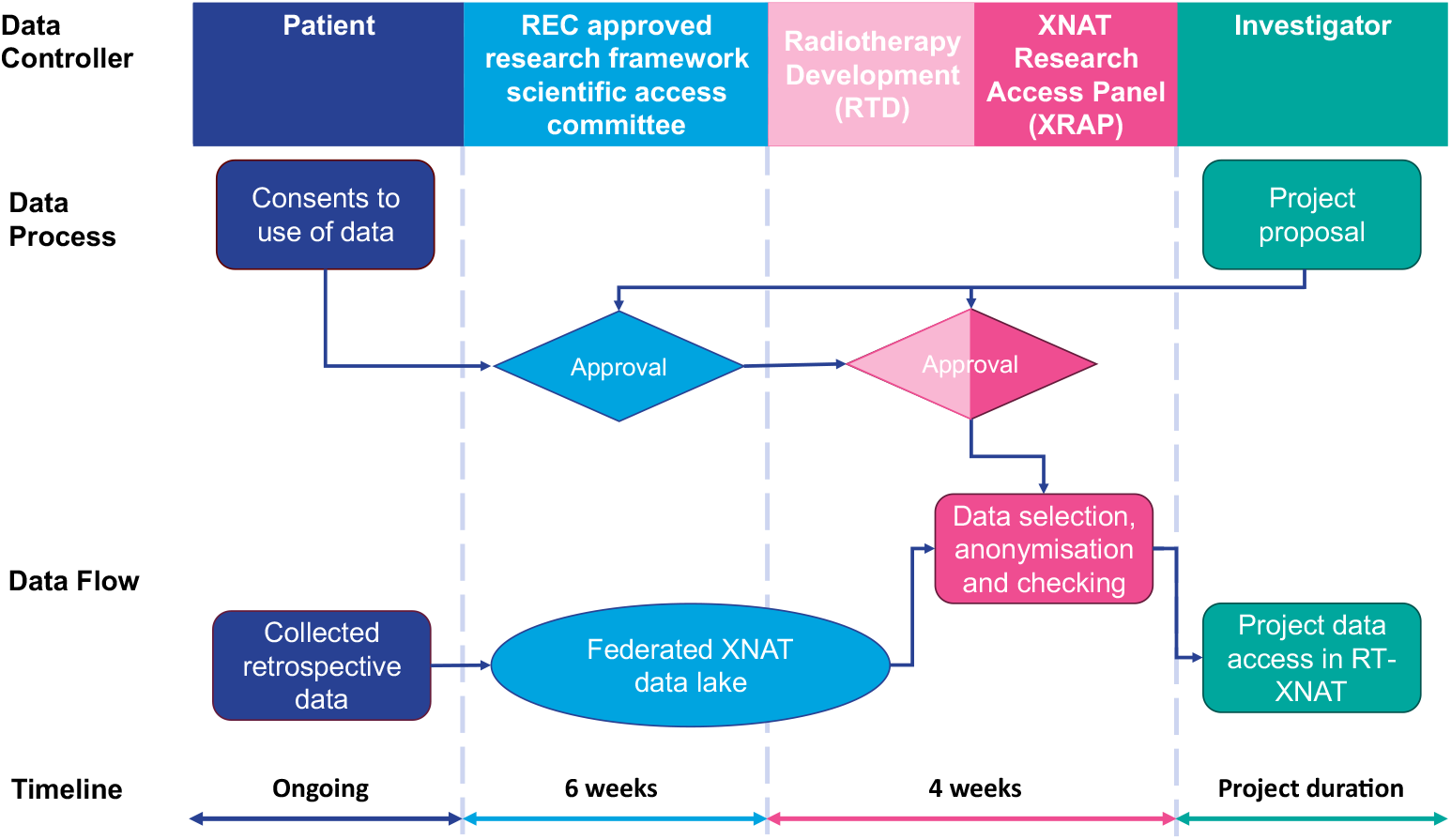
Research data access pipeline with data controllers and anticipated timeframes displayed.

### Imaging Data

Over 80,000 imaging studies with accession numbers were located within PACS comprising of 834 study descriptions; 115 were pertinent to HNC diagnosis, staging, and monitoring (Supplementary Data: Appendix 1). Non-relevant studies included over 10,000 ultrasounds, 27,000 planar x-rays and others (e.g. barium swallows, mammograms, angiograms, DEXA scans). This led to the attempted ingestion of over 35,000 studies into XNAT. 20,726 were successfully received by XNAT using the REST-API. 12,054 accession numbers lacked imaging (e.g. ^18^F-FDG PET-CT injections). An audit of a subset (5%) of 3,163 failures found that 100% of CTs and MRIs were either cancelled or had images under a different accession number, already ingested. For ^18^F-FDG PET-CT scans, VF and XR studies, the majority had been cancelled or no images were available in PACS; but 20% of cases had images XNAT couldn’t retrieve. An imaging summary .csv file flags missing images for manual retrieval if needed, ensuring data completeness. The multimodal dataset is shown in Figure 3.

**Figure 3:**
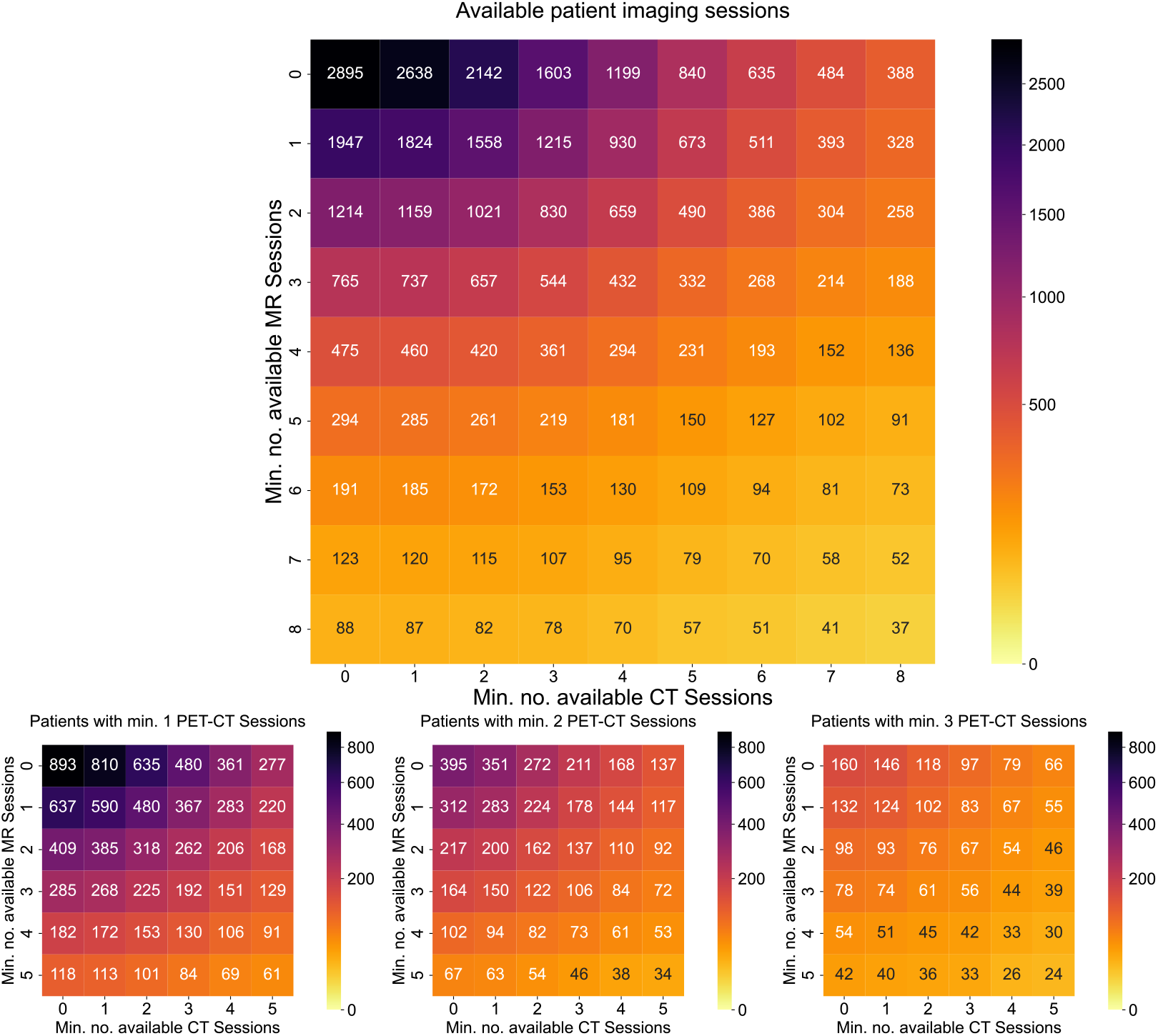
Multimodal nature of minimum number of imaging sessions available per patient.

### Radiotherapy Data

Figure 4 shows available RT datasets. Of 2,895 patients, 2,581 patients received HNC RT, 314 never did and 166 patients were initially treated before inversely planned IMRT, leaving 2170 patients for inclusion. Most excluded radical treatment cases were T1 glottic larynx patients (n=84).

**Figure 4:**
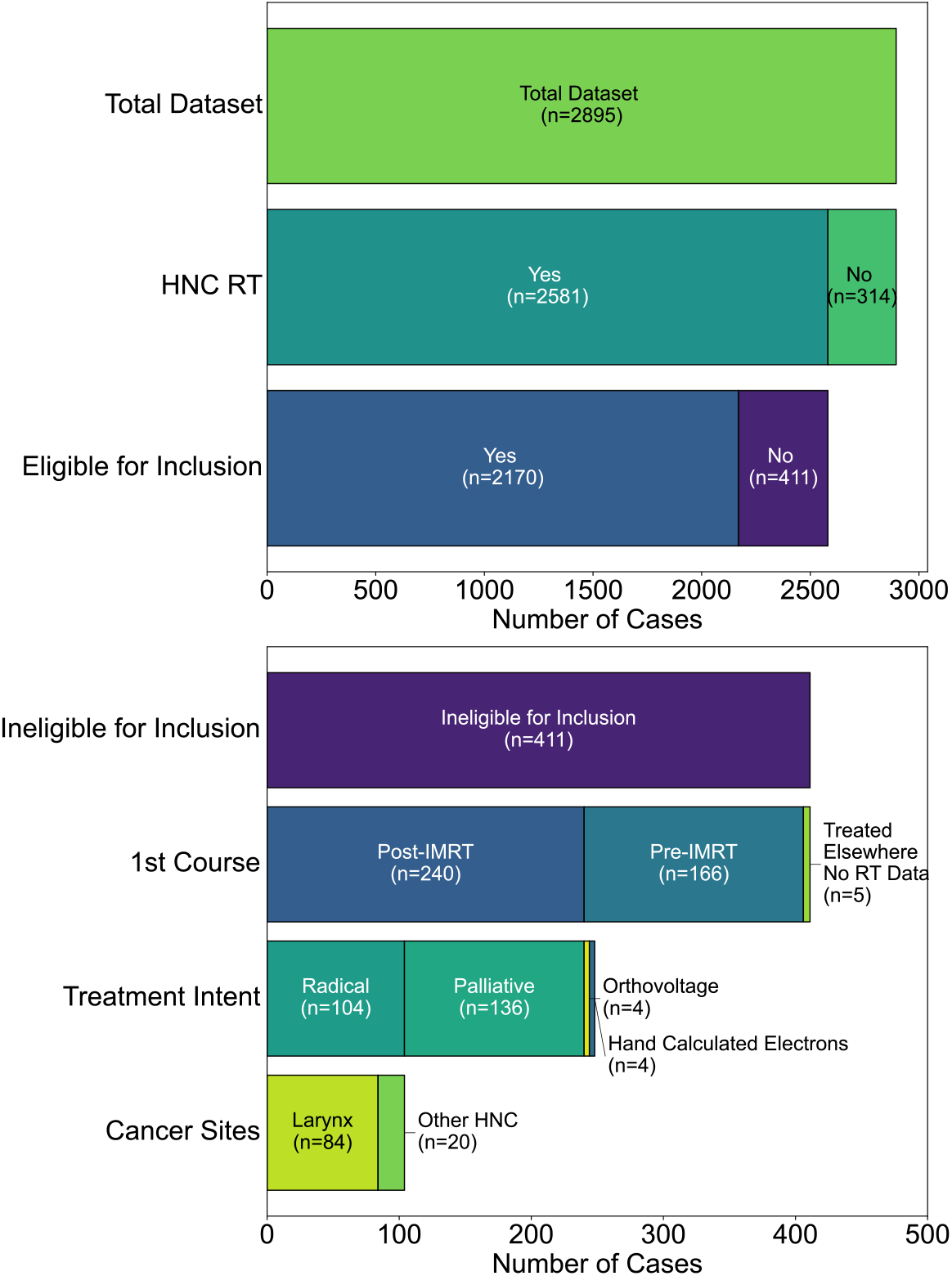
Figure of eligibility of inclusion of RT datasets within XNAT datalake. The majority of cases are excluded due to their reduced relevance to treatment techniques today e.g. before IMRT was introduced at the centre or to the treatment site (Larynx).

From the legacy TPS Monaco, 932 patients were included but retrieval issues arose. Early IMRT patients had dose cube misalignments in Eclipse and a 2016 Monaco database corruption complicated plan reconstruction. Overall, 2,071 patients had their RT datasets completely retrieved into XNAT. The remaining 99 patients had known data quality issues, which can be summarised as: dose cube misalignment (n=24), having missing full datasets (n=52) and having partial missing datasets (n=23) where the patient was known to have been replanned but either the replan or the original plan could not be located.

Figure 5 shows data availability over time. Peaks correspond to early IMRT dose misalignments and 2015-2016 database corruption. No issues were found in Eclipse-era datasets but COVID-19’s impact is visible in 2020 patient numbers, in line with pan-London reduced referral rates [25].

**Figure 5:**
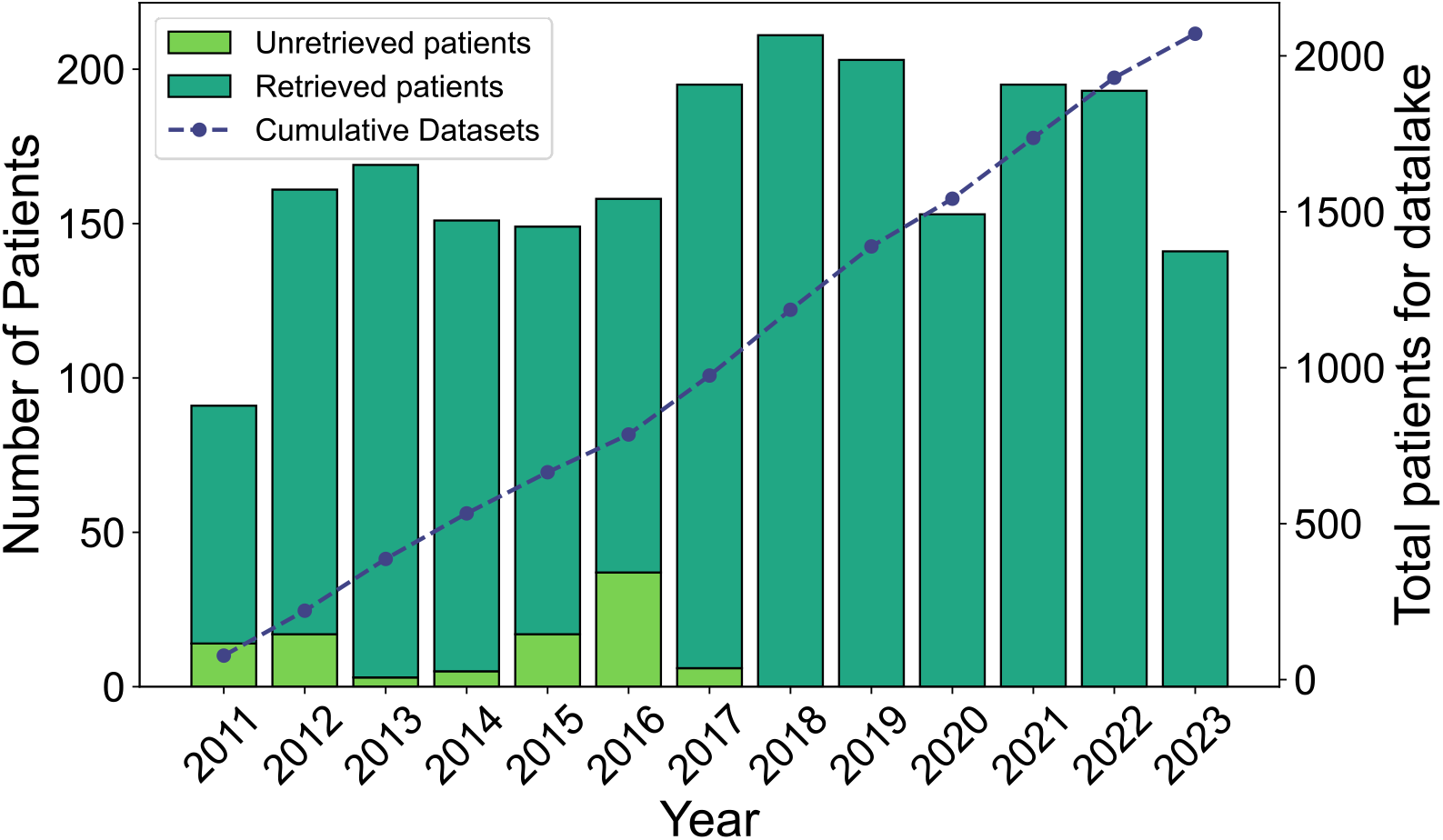
Data lake growth of eligible RT patients with retrieved data displayed. Year 2023 reflects January to September cases.

### Clinical Data

Data was collected for 2,895 patients covering demographics, comorbidities, diagnosis, treatment and follow-up. The dataset contains 1,803,670 completed data points including ethnicity, alcohol intake, smoking status, staging, relevant blood tests, chemotherapy details, RT prescriptions, survival and toxicity data. A data dictionary of 2,077 categories was compiled. An additional 4,212,494 data points were marked as Unknown/NULL/Not stated, often due to expected absences (e.g. 2-year toxicity data for a patient who died within a year) rather than true ‘missing’ data not yet curated.

### Recommendations

Table 1 displays a summary of the technical and practical recommendations for creating a high-quality retrospective cohort database and controlling the access to the data.

**Table 1:** Summary of recommendations to construct a retrospective cohort clinically annotated imaging and radiotherapy database.

|  |  |
| --- | --- |
| 1. Patients' rights and data protection | The utmost care should be taken to include only patients who have given explicit informed consent to have their data used in research. Users of the database should have undergone basic training in data management and GCP (Good Clinical Practice) in research. Any data used in research should be fully anonymised. |
| 2. Patient and data definition | Strict eligibility criteria for cohort definition and data admission should be employed to adhere to best practice data minimisation principles. Protocols for data collection harmonisation should be established to facilitate research studies and institutional and cross-institutional collaborations. Attention should be paid to the accuracy, precision and consistency of the data. Sourcing data from multiple places and comparing the outputs increases the reliability of the data. Known missing data should be labelled for completeness. |
| 3. Infrastructure | The longevity of the technical solution should be considered including scalability, maintenance, software upgrades, safe and sustainable network communications, accessibility of data, resource and ownership implications. |
| 4. Multi-Disciplinary Team buy-in | Both clinical and technical skills are required to construct a database with high quality and clinically annotated radiotherapy and imaging data. |
| 5. Legacy data | The relevance of legacy data to relevant and current practice and the scarcity of data with comparison to the length of retrieval should be considered. |

## Discussion

The data lake constructed in XNAT represents a comprehensive repository of imaging and associated clinical data for 2895 HNC patients treated by the clinical oncology team at our organisation who received radiotherapy as part of their treatment. The retrospective nature of the database enables the inclusion of real-world variability which is particularly valuable for studying differences in treatment response and the cohort size enables robust subgroup analyses and more generalisable findings.

Existing HNC databases vary in scope and focus. Our repository bridges gaps often observed in conventional databases through the combination of clinical, demographic, imaging and treatment planning data in a single governed entity. In the UK, existing datasets provide valuable insights into specific aspects of patient care such as the Radiotherapy Dataset (RTDS) [26] capturing comprehensive data on RT treatments in England or the Head and Neck Cancer Audit (HANA) [27] collecting diagnosis, staging, treatment and pathway information but not including imaging or detailed clinical outcomes which limits its utility for holistic research approaches, including multimodal AI.

This trend of databases catering to either in-depth clinical information or imaging information with sparse clinical data is reflected worldwide. The National Cancer Institute’s TCIA (The Cancer Imaging Archive) emphasises imaging datasets for machine learning applications however many existing resources lack the depth and breadth and integration of multimodal data that this data lake offers. By containing CT, PET-CT and MRI images as well as RT doses and images, the breadth of our imaging data lake surpasses that of data contained by Head-Neck-Radiomics-HN1 [28], HNSCC [29][30], Head-Neck-PET-CT [31] and RADCURE [32][33]. The most comparable open-source dataset of its size available worldwide is RADCURE [32][33] which contains 3346 patients and contains radiotherapy images and contours. Our radiotherapy dataset is deeper in scope containing dose cubes to enable prognostic modelling leveraging dosimetric factors. Additionally, we have multimodal radiotherapy imaging beyond just CT images. These additional imaging modalities may provide information beyond patients’ clinical indicators and their disease characteristics beyond the anatomical and density information provided by CT. Additionally, no datasets currently on TCIA contain dental x-rays. RADCURE does however have standardised the nomenclature for individual organ at risk and GTV contours which would be a valuable extension in scope for our group in the future. Due to the ethics approval requirements, our database cannot be made open source, but GCC welcomes specific and detailed proposals for new collaborations [20], including external organisations, subject to appropriate governance approvals including Data Sharing Agreements.

The optimal performance of machine learning models and their generalizability is hinged on the quality of data used for model construction. Throughout the construction and validation phases, care was taken to respect the data quality dimensions set out by regulatory bodies including NICE [24] to ensure reliable and relevant data of robust quality to enable data use for meaningful RWE generation [7]. By undertaking the dataset validation and verification checks, we have demonstrated the accuracy, completeness and consistency of our dataset across critical variables and prevented creation of a data dump/swamp [6]. There are still known deficits in missing data due to corrupted datasets from legacy systems and difficulty of retrieval. By annotating these deficits, we hope that future researchers can evaluate for themselves whether it is worth the additional time and expense involved in retrieving this data. These issues highlight the complexity in creating a unified database from legacy systems. The experience also demonstrates the need to consider the ease of data retrieval and system interoperability when procuring new technical systems in order to reduce future data management challenges and support continuity of care and research over time.

We have created a secure and extensible imaging and H&N RT cancer database. This data lake for H&N research will facilitate much quicker data access for future AI and research projects in H&N RT. The secondary XNAT provides safeguards for data access and control, securing the data and ensuring it is used in accordance with high standards of information governance. Currently work is underway to update our database with new events and cases occurring since October 2023. The integration of imaging and clinical data in a standardised format enhances both the accessibility and reusability of the data [14]. This database set-up promises to be an exceedingly useful tool for research, revolutionising the time and cost associated with the production and validation of machine learning models. We have shared our governance structure and hope it will be useful for other UK and worldwide providers to produce a similar research and audit tool.

We have also provided a list of recommendations that we believe are vital to create a high-quality retrospective cohort database that adheres to the highest standards of patient dignity and respect as well as preserving and enhancing the usefulness of routine data generated during standard quality of care of cancer patients.

## Data Availability

All data produced in the present work are contained in the manuscript.

## Acknowledgements

We would like to thank the patients of Guy’s Cancer for their support of this database. We would also like to thank the Information Governance team at Guy’s and St. Thomas’ NHS Foundation Trust for supporting this work.

## Supplementary Material

**Figure S1:**
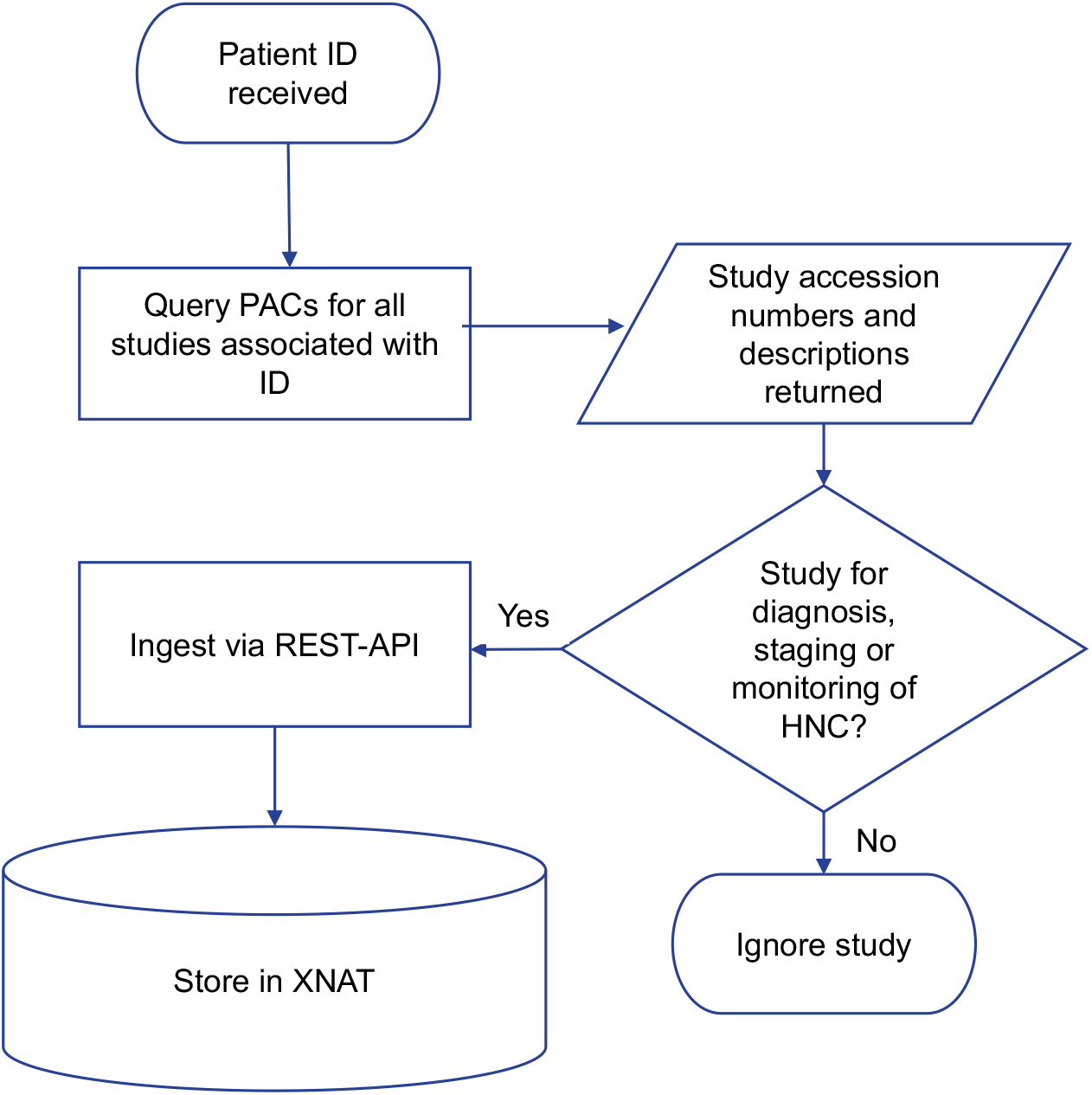
Imaging data flow from PACs to XNAT.

**Figure S2:**
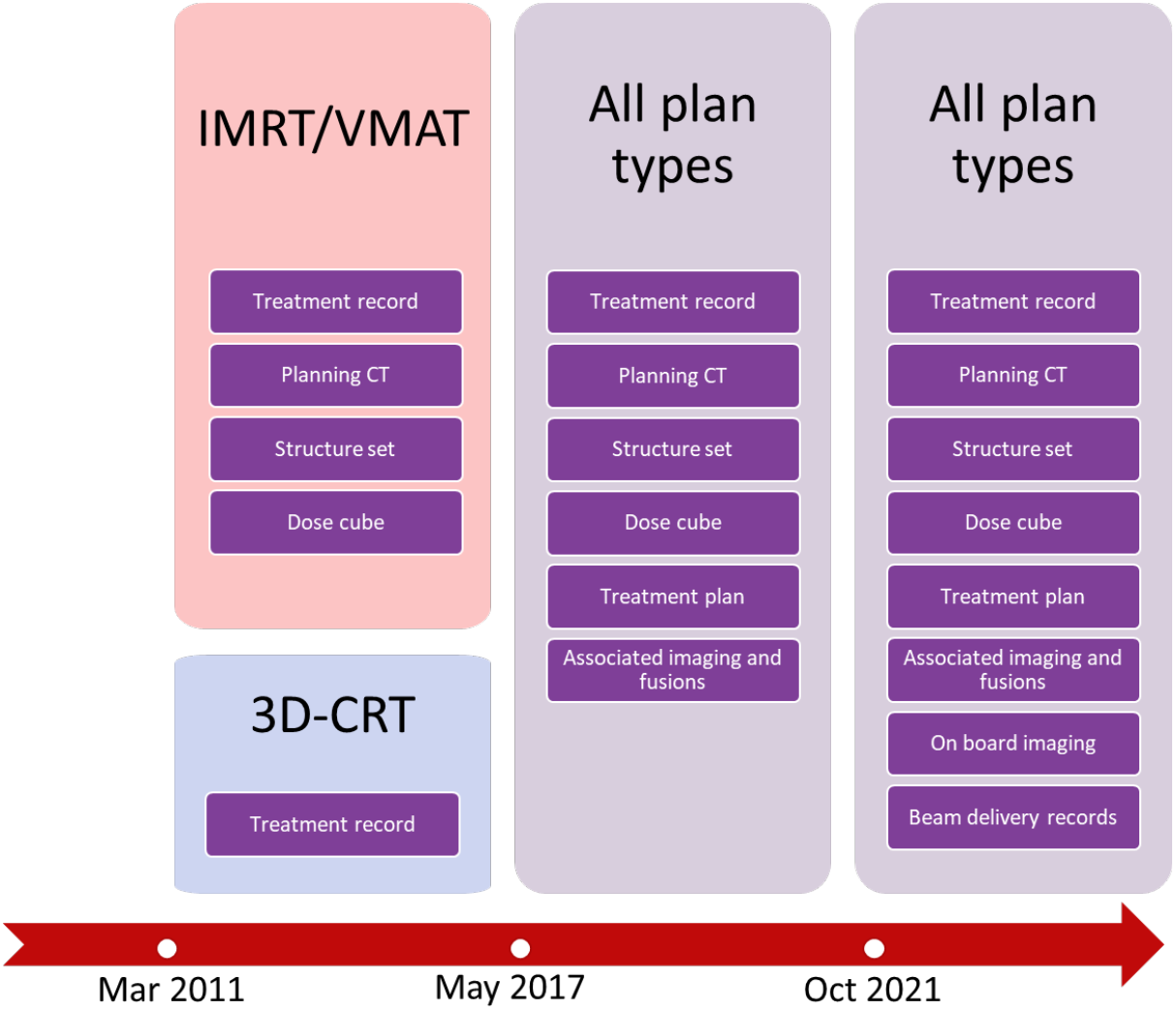
Data retrieved from each radiotherapy epoch. Between 2011-2017, 3D-CRT plans were created in XiO and IMRT/VMAT plans were produced within Monaco.

**Figure S3:**
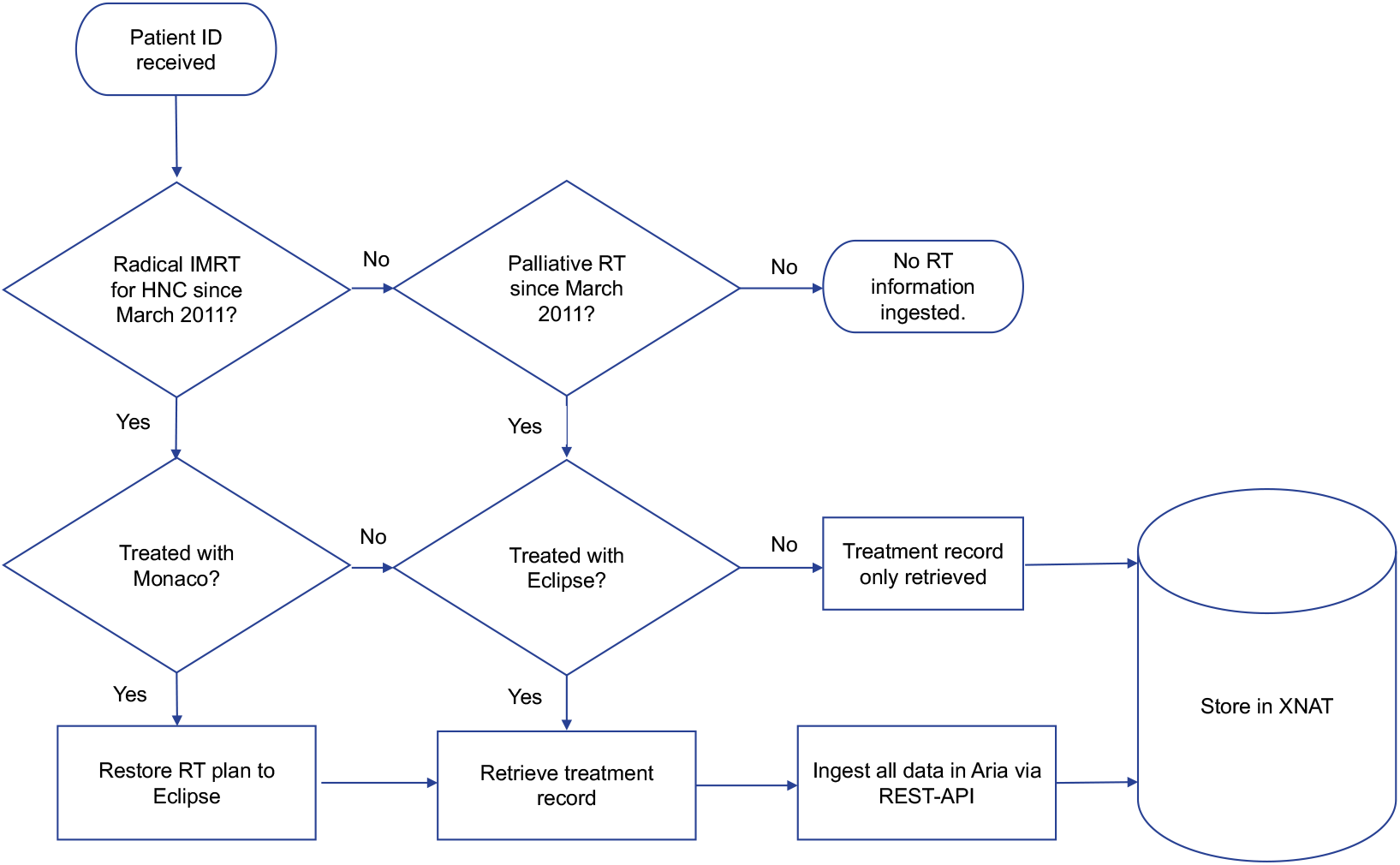
Radiotherapy data flow from Radiotherapy systems to XNAT.

## Appendix 1: Retained imaging study descriptions

|  |  |  |
| --- | --- | --- |
| Brain - MRI | CT Thorax & abdomen | MRI Thorax |
| CT Abdomen & pelvis with contrast | CT Thorax & abdomen & pelvis | MRI Thorax with contrast |
| CT Abdomen and pelvis with contrast | CT Thorax & abdomen with contrast | MRI Trigeminal nerve |
| CT Adrenal with contrast Both | CT Thorax abdo pelvis with contrast | MRI Whole body |
| CT chest high resolution with contrast | CT Thorax abdomen and pelvis | Neck - CT |
| CT Chest HRCT | CT Thorax abdomen pelvis with contrast | NM FIGARO FDG PET CT |
| CT Chest/Abdomen/Pelvis with Contrast | CT Thorax and abdomen | NM Half body FDG PET CT |
| CT Face | CT Thorax and abdomen with contrast | NM Halfbody + H&N FDG PET CT |
| CT Face with contrast | CT Thorax with contrast | NM Halfbody FDG PET CT |
| CT Facial bones | DENTAL-UD Craniofacial Soft Tissues | NM INGRES FDG PET CT |
| CT Head | Face - CT | NM Javelin FDG PET CT |
| CT Head with contrast | Mandible PA - Dental | NM PET-MR in HNC RT pilot PET MR |
| CT IAM with contrast | MRI - add Gadolinium | NM PET-MR Role in Oncology |
| CT Internal auditory meatus Both | MRI Adrenal Both | NM Whole body FDG PET C |
| CT Liver | MRI Face with contrast | NM Whole body FDG PET CT |
| CT Liver triple phase | MRI Head | OPG - X-Ray |
| CT Liver with contrast | MRI Internal auditory meatus Both | PET CT Scan |
| CT Mandible | MRI Larynx | Research - MRI (specify) |
| CT Mastoid Both | MRI Liver & Spleen | Unspecified CR NECK |
| CT Neck | MRI Liver and spleen | Unspecified CT CHEST |
| CT Neck & thorax | MRI Liver with hepatobiliary contrast | Unspecified CT CHEST/ABDO/PELV |
| CT Neck & thorax & abdo & pelvis & cont | MRI Neck | Unspecified CT HEAD |
| CT Neck & thorax & abdomen | MRI Neck with Gadolinium | Video swallow |
| CT Neck & thorax & abdomen & contrast | MRI Orbit Both | Video/fluoroscopy Speech |
| CT Neck & thorax with contrast | MRI Perfusion weighted | XR Dental bitewing |
| CT Neck and thorax with contrast | MRI Pituitary | XR Dental mandibular occlusal |
| CT Neck thorax abdomen pelvis with Cont | MRI Radiotherapy planning scan head | XR Dental maxillary occlusal |
| CT Neck with contrast | MRI Radiotherapy planning scan neck | XR Dental oblique lateral Both |
| CT Orbit Both | MRI Radiotherapy planning scan pelvis | XR Dental oblique lateral Lt |
| CT Orbit with contrast Both | MRI Salivary glands | XR Dental oblique lateral Rt |
| CT Pituitary with contrast | MRI Sinuses | XR Dental periapical 10 |
| CT Planning scan | MRI Spinal Cord | XR Dental periapical 10+ |
| CT plus Contrast | MRI Spine cervical | XR Dental periapical 5 |
| CT Radiotherapy planning scan | MRI Spine lumbar & sacral | XR Dental periapical full |
| CT RT PLANNING | MRI Spine lumbar and sacral | XR Orthopantomogram full |
| CT Sinuses | MRI Spine thoracic | XR Orthopantomogram Lt |
| CT Sinuses with contrast | MRI Spine whole | XR Orthopantomogram Rt |
| CT Thorax | MRI Spine whole with contrast |  |
| CT Thorax & abdo & pelvis with contrast | MRI Temporomandibular joint Both |  |

